# Nutritional availability and carbon footprints of vegetarian and vegan diets: a cross-sectional analysis of dietary data for UK children

**DOI:** 10.64898/2026.01.28.26345075

**Authors:** Alice Coffey, Robert Lillywhite, Oyinlola Oyebode

## Abstract

As plant-based (PB) diets become more common among UK children, understanding their nutritional adequacy and environmental impact is vital. This study assessed nutrient intake and dietary greenhouse gas emissions among children following omnivorous, vegetarian, and vegan diets.

A cross-sectional analysis was conducted using three-day weighed food diaries from 39 UK children aged 2–12 years (omnivore n=15; and PB: vegetarian n=11; vegan n=13). Nutrients were analysed with and without supplementation using Nutritics software. GHGEs were calculated at the ingredient level (kgCO₂e/day) and grouped by Eatwell Guide food categories.

No dietary group met all nutrient reference values. Omnivores exceeded recommended intakes for saturated fat and free sugars while failing to meet the recommended intake for fibre, whereas PB children had intakes of these nutrients in the healthy range. PB diets were adequate in protein and vitamin B12 even in the absence of supplementation. Vegan children also met iron requirements from diet alone, whereas omnivore and vegetarian children did not meet iron targets without supplementation. Vitamin D intake was insufficient across all groups when supplements were excluded, with only vegan children achieving recommended levels through supplementation. Zinc requirements were met only by vegetarian children with the aid of supplements and were not met by vegan or omnivore children with or without supplementation. Iodine intake remained inadequate in vegan children even with supplementation. Mean daily GHGEs differed significantly between diet groups (p < 0.001): omnivores having the highest emissions, while vegans had the lowest emissions: 46% lower than omnivores, and 20% lower than vegetarians.

Well-planned PB diets can meet most nutrient needs in UK children when supported by fortified foods and supplements, while substantially reducing dietary GHGEs compared with omnivorous diets. Shifting away from animal protein and dairy provides the greatest opportunity for improving both nutritional quality and environmental sustainability.

## 1. Introduction

Interest in plant-based (PB) eating has increased in recent years, with nearly half of UK residents reporting efforts to reduce their meat consumption (1). Although many of these individuals still consume some animal products, this shift shows a move towards more vegetarian and vegan diets. Although there is no robust evidence of prevalence, an online poll completed by YouGov found that 2% of the UK population identified as vegan (diets containing no animal products) and 6% as vegetarian (diets containing no meat or fish); for those aged 18-24 this rose to 5% and 11%, respectively (2). While a growing body of evidence supports the nutritional adequacy for more PB diets (used here as a combined term for vegetarian and vegan diets) in adults (3,4), there is limited evidence of their nutritional adequacy in children, particularly in the UK context.

As the adoption of PB diets is increasing, it is essential to understand if these diets can meet children’s nutritional needs. This is of particular importance in light of the “double burden of malnutrition” that many high-income countries, including the UK are now facing in children, with rising obesity rates and increasing micronutrient deficiencies (5). Data from the National Diet and Nutrition Survey (NDNS), shows children in the UK are at risk of deficiency in key nutrients, in particular, vitamin D, fibre, folate and iron, while exceeding recommendations for free sugars and saturated fat (6). These findings reinforce the need to evaluate if PB diets can provide adequate nutrition for children without adverse effects.

Some studies have highlighted potential concerns with PB diets in children, a recent systematic review including 30 global studies with children aged 2-18, reported that children following PB diets may be at risk of deficiencies in iron, zinc and B12 (7). Additionally, both PB children and omnivore children were at risk of deficiency of vitamin D, calcium and iodine (7). However, a recent cross-sectional study undertaken in Germany, with 820 participants aged 1-18 showed that PB diets offer some health benefits and are higher in fibre and lower in saturated fat and free sugars, compared with omnivore diets (8). Diets high in red and processed meats have been linked to increased risk of cardiovascular disease, certain cancers and type 2 diabetes (T2D) (9). It has been shown that children consuming vegan diets have healthier cardiovascular risk profiles (10), but the long-term evidence remains unexplored.

This study aims to address the critical gap in the literature by analysing the nutritional composition and carbon footprints of omnivore, vegetarian and vegan diets in UK children aged 2-12.

## 2. Methodology

### 2.2 Study design and participants

This study is a cross-sectional study of dietary data from UK children. Participants were recruited online via convenience and snowball sampling, with the electronic invitation circulated on social media between March 2023 and November 2023. Inclusion criteria were those with omnivore, vegetarian or vegan children, between the ages of 2 and 12 years, living in the UK. Excluded participants were those who only had children outside the 2-12 age range in their care, or who lived outside the UK, those who followed a different diet to vegan, vegetarian or omnivore (e.g. pescatarian) and those with medical conditions that caused them to have a highly restrictive diet. Parents participating with their children received a £10 shopping voucher for taking part, this amount was chosen to encourage participation, without being considered a bribe. Nutritional outcomes were also shared with parents. Research ethics approval was obtained from the University of Warwick’s Biomedical Sciences Research Ethics Committee (BSREC). Application reference: **BSREC 76/22-23**

### 2.3 Sample size

The sample size for this study was restricted to 15-45 children based on available resources, including funding and time. There are no UK studies that compare vegan, vegetarian and omnivore diets in children; however, it is common for weighed dietary studies to have similar sample sizes to this to give indicative results (11–13).

### 2.4 Nutrition assessment

Dietary intake was assessed using three-day weighed food diaries, completed by parents of the participating children through the Qualtrics survey software online. Electronic kitchen scales and instructions were provided for parents to weigh all food and drink that their child was served. Leftover food was also recorded, using fraction estimations – e.g. half the broccoli left. For school lunches, where parents could not weigh the food, meals were recorded and study staff estimated quantities using the school food plan portion guide as a reference. Parents also asked children if they left any food. Breast milk was recorded through minutes fed, with average amounts per minute used to estimate quantities. Parents were asked to provide brands of food, drinks and supplements consumed to allow for more accurate analysis. Where these weren’t provided, the most popular product from the Nutritics database were used, for plant milks, these were a fortified version. Dietary data were input into Nutritics software (14) and nutrition reports were generated.

### 2.5 Demographic data

Demographic data were collected online using Qualtrics. Demographic questions asked were:

- Relation to child participating
- Postcode (First part only, e.g. CV4)
- Household income bracket
- Qualifications
- Marital status

Some non-identifiable data about the participating children was also collected:

- Child’s sex
- Age
- Ethnicity
- School lunch status
- What type of school does the child go to (private, state etc.)
- What dietary pattern does the child follow

The demographic data collected helped identify trends and associations. Parents were asked the sex of their child, rather than gender, to compare with dietary reference values set for male and females.

### 2.6 Data analysis

#### 2.6.1 Demographic characteristics

The demographic data collected were analysed by dietary pattern. This provided insights into the characteristics and distribution of the participants within each dietary group. Statistical analysis of the demographic variables was conducted using SPSS software (15). Analysis of Variance (ANOVA) tests were performed to determine the significance of differences in characteristics between the groups with continuous data (age and index of multiple deprivation) and chi-squared used for categorical data (child ethnicity, school type, lunch type, child sex, parental qualifications, marital status, income group), with post-hoc tests conducted to see significant differences between dietary patterns.

#### 2.6.2 Nutrients

The collected dietary data was input into the Nutritics software (version 6.14), where macro and micronutrient data were computed. These nutrients included calories, total fat, saturated fat, protein, carbohydrate, fibre, free sugar, vitamins A, C, B12, and D, sodium, iodine, iron, calcium, zinc, and folate. These micronutrients were identified as potential causes for concern in past papers and government data (Alexy et al., 2021; Desmond et al., 2021; Office for Health Improvement & Disparities, 2025). These results were then input into SPSS (15), where they underwent statistical analysis. For each dietary pattern, the mean value for each nutrient was calculated. ANOVA analysis was performed to identify statistically significant differences in nutrient levels between groups. This analysis was conducted on data both including and excluding dietary supplements to observe the impact of supplementation. After calculating the overall mean for each nutrient, a more detailed analysis was carried out. Mean nutrient intakes for each dietary pattern were compared with the Department of Health’s dietary reference guidelines for children (16). Given that nutrient recommendations vary by age and sex, adjustments were made accordingly, and statistical analysis determined the percentage of each nutrient recommendation met. This provided the mean percentage of nutrients met for each dietary pattern, examined with and without supplements.

#### 2.6.3 Carbon footprint analysis

Based on the dietary data collected, detailed product and ingredient lists were generated for each dietary pattern, identifying exactly what was consumed by each participant. Carbon footprint values were then assigned at the ingredient level. Where available, values were sourced from established databases (primarily CarbonCloud and LiveLCA) (17,18), with preference given to UK-specific data or the closest geographical equivalent.

For ingredients lacking direct carbon data, estimates were derived using the closest comparable product, supported by expert judgement. For example, in the absence of a value for a vegan croissant, the footprint of a standard butter-based croissant was used, with a 50% reduction applied to account for the absence of dairy fat. Where no suitable proxy was available, carbon footprints were calculated from the ingredient composition. For instance, the carbon footprint of a ‘Yoyo Bear’ fruit snack (65.8% apple, 32.9% pear, 1% strawberry, 0.3% black carrot extract) was estimated using weighted averages for apples and pears.

All carbon values were standardised to kgCO₂e per 100 g to ensure comparability. These were then scaled to reflect the actual quantities consumed by each child, as recorded in the three-day weighed food diaries. This enabled the calculation of each participant’s daily dietary carbon footprint, from which mean values were derived across the study period.

Individual dietary data were then grouped into the Eatwell Guide food categories: fruit and vegetables; starchy foods/carbohydrates; protein (including meat, fish, pulses and PB alternatives); dairy (including dairy alternatives); fats (oils and spreads); and HFSS foods (crisps, biscuits, cakes, condiments). A further ‘other’ category was created for items not captured by the above (e.g. vinegar, spices). Composite foods were allocated according to their primary ingredient (e.g. margarita pizza to carbohydrate, pesto to oils and spreads).

Daily mean greenhouse gas emissions (GHGEs) were calculated for each individual across these categories. Overall group means were then compared using one-way ANOVA with post-hoc tests conducted in SPSS to assess differences between dietary patterns. Eatwell Guide categories were chosen to align the analysis with national dietary recommendations and to identify which food groups contribute most to dietary GHGEs in different dietary patterns, thereby supporting policy-relevant interpretation.

## 3. Results

### 3.1 Demographics

The study population was comprised of 39 participants: 15 omnivores, 11 vegetarians, and 13 vegans, providing balanced groups for reliable assessment (Table 1). Most demographic characteristics did not differ significantly between dietary patterns. Across all groups, the majority of participants were male (>60% in each), though the distribution of sex was not statistically different between groups.

**Table 1:** Demographic characteristics of study participants.

| Diet Pattern<br>(overall % of participants) | Vegan (33%) | Vegetarian<br>(28%) | Omnivore<br>(39%) | P-Value |
| --- | --- | --- | --- | --- |
| Study Size 39 | 13 | 11 | 15 | n/a |
| Supplement use |  |  |  |  |
| Yes | 12 (92%) | 9 (82%) | 2 (13%) | <0.001 |
| No | 1(8%) | 2 (18%) | 13 (87%) |  |
| Sex |  |  |  |  |
| Female | 4 (31%) | 4 (36%) | 6 (40%) | 0.878 |
| Male | 8 (69%) | 7 (64%) | 9 (60%) |  |
| Age (years) | Mean= 5.2<br>(std.dev=2.68<br>) | Mean= 5.73<br>(std.dev=2.72<br>) | Mean= 5.23<br>(std.dev=2.80<br>) |  |
| 2 | 3 (23%) | 1 (9%) | 2 (13%) | 0.870 |
| 3 | 2 (15%) | 2 (18%) | 4 (27%) |  |
| 4 | 0 (0%) | 1 (9%) | 1 (7%) |  |
| 5 | 2 (15%) | 1 (9%) | 2 (13%) |  |
| 6 | 2 (15%) | 3 (27%) | 1 (7%) |  |
| 7 | 1 (8%) | 0 (0%) | 2 (13%) |  |
| 8 | 2 (15%) | 1 (9%) | 0 (0%) |  |
| 9 | 0 (0%) | 0 (0%) | 2 (13%) |  |
| 10 | 0 (0%) | 2 (18%) | 1 (7%) |  |
| 11 | 1 (8%) | 0 (0%) | 0 (0%) |  |
| Ethnicity (n) |  |  |  |  |
| White | 13 (100%) | 10 (91%) | 11 (74%) | 0.099 |
| Non-white | 0 (0%) | 1 (9%) | 4 (26%) |  |
| School Type (n) |  |  |  |  |
| State School (non-selective) | 5 (39%) | 8 (73%) | 8 (53%) | 0.200 |
| Home Schooled/Stay at home | 2 (15%) | 0 (0%) | 1 (7%) |  |
| Flexi school: Part home schooled/part school | 2 (15%) | 0 (0%) | 0 (0%) |  |
| Private School | 0 | 0 (0%) | 1 (7%) |  |
| Nursery | 3 (23%) | 3 (27%) | 5 (33%) |  |
| Childminder | 1 (8%) | 0 (0%) | 0 (0%) |  |
| School lunch type (n) |  |  |  |  |
| Packed Lunch | 5 (39%) | 1 (9%) | 0 (0%) | 0.007 |
| School Provided Lunch | 0 | 7 (64%) | 11 (73%) |  |
| Mix: School Provided or Packed | 5 (39%) | 3 (27%) | 3 (20%) |  |
| At Home | 2 (15%) | 0 (0%) | 1 (7%) |  |
| Missing data | 1 (8%) | 0 (0%) | 0 (0%) |  |
| Index of Multiple Deprivation |  |  |  |  |
| Unavailable | 4 (31%) | 2 (18%) | 0 (0%) | 0.052 |
| 1 | 0 (0%) | 0 (0%) | 0 (0%) |  |
| 2 | 0 (0%) | 0 (0%) | 0 (0%) |  |
| 3 | 1 (8%) | 0 (0%) | 0 (0%) |  |
| 4 | 2 (15%) | 3 (27%) | 1 (7%) |  |
| 5 | 0 (0%) | 1 (9%) | 4 (27%) |  |
| 6 | 2 (15%) | 1 (9%) | 1 (7%) |  |
| 7 | 1 (8%) | 0 (0%) | 0 (0%) |  |
| 8 | 1 (8%) | 1 (9%) | 6 (40%) |  |
| 9 | 1 (8%) | 3 (27%) | 0 (0%) |  |
| 10 | 1 (8%) | 0 (0%) | 3 (20%) |  |
| Household<br>Income |  |  |  |  |
| Less than £20,000 | 1 (8%) | 1 (9%) | 0 (0%) | <0.001 |
| £20,000 - £40,000 | 2 (15%) | 1 (9%) | 0 (0%) |  |
| £40,001 - £60,000 | 8 (60%) | 1 (9%) | 1 (7%) |  |
| £60,001 - £80,000 | 2 (15%) | 5 (46%) | 3 (20%) |  |
| Over £80,000 | 0 (0%) | 3 (27%) | 11 (73%) |  |
| Parental marital<br>Status |  |  |  |  |
| Married/same sex<br>civil partnership | 9 (69%) | 7 (64%) | 15 (100%) | 0.082 |
| Cohabiting | 4 (31%) | 3 (27%) | 0 (0%) |  |
| Divorced/separate<br>d | 0 (0%) | 1 (9%) | 0 (0%) |  |
| Highest Parental<br>Qualification |  |  |  |  |
| No qualifications |  |  |  | 0.758 |
| Level 1: one to four GCSE passes (grade A* to C or grade 4 and above) and any other GCSEs at other grades, or equivalent qualifications | 0 | 0 | 0 |  |
| Level 2: five or more GCSE passes (grade A* to C or grade 4 and above) or equivalent qualifications | 0 | 0 | 0 |  |
| Apprenticeships | 0 | 0 | 0 |  |
| Level 3: two or more A Levels or equivalent qualifications | 0 | 1 (9%) | 1 (7%) |  |
| Level 4 or above: Higher National Certificate, Higher National Diploma, | 12 (92%) | 8 (73%) | 12 (80%) |  |
| Bachelor's degree,<br>or post-graduate<br>qualifications |  |  |  |  |
| Other<br>qualifications, of<br>unknown level | 1 (8%) | 2 (18%) | 2 (13%) |  |

Mean age was comparable, with omnivores averaging 5.2 years, vegetarians 5.7 years, and vegans 5.2 years; these differences were not statistically significant (p=0.870). Ethnicity distribution also did not differ significantly (p=0.099), although most participants identified as white (74% of omnivores, 91% of vegetarians, and 100% of vegans). School type showed no significant variation across groups. There was a significant difference in school lunch type between dietary groups (p= 0.007). However, post-hoc comparisons with correction for multiple testing did not reveal significant differences between any specific groups. No significant differences were observed in index of multiple deprivation, parental education level, or marital status. Household income differed significantly between dietary patterns overall (p=< 0.001). Post-hoc tests indicated that omnivores reported significantly higher household income than vegans, while no significant differences were observed between the other groups. Although overall differences in parental education level were not statistically significant, most parents were educated to degree level or higher (omnivores 100%, vegetarians 91%, vegans 93%).

Analysis of supplement use revealed clear differences between dietary patterns. Almost all vegans (92%) and most vegetarians (82%) reported supplement use, compared with only 13% of omnivores. A chi-square test indicated a significant association between diet type and supplement use (p=< 0.001). Post-hoc comparisons showed that supplement use was significantly more common among both vegans and vegetarians compared with omnivores, while no significant difference was observed between vegans and vegetarians.

### 3.2 Dietary Intakes

Table 2 shows the mean percentage of nutrient intake requirements met for each diet group. This has been adjusted for age and sex, where intake recommendations differ. Nutrients were calculated with and without nutrient supplements. When observing each individual diet, omnivores do not meet the recommended intake for calories, total fat, carbohydrate, fibre, zinc and vit D and exceed recommendations for saturated fat, protein, free sugar, vitamin b12, vitamin A, vitamin C, Sodium, Iodine, Iron, Calcium and folate, when supplements are included. When excluding supplements, there is a decrease in vitamin B12, A, C, and folate, however, they still all exceed the recommended intake. Iron, zinc and vitamin D also decrease, not reaching recommended intakes.

**Table 2:**
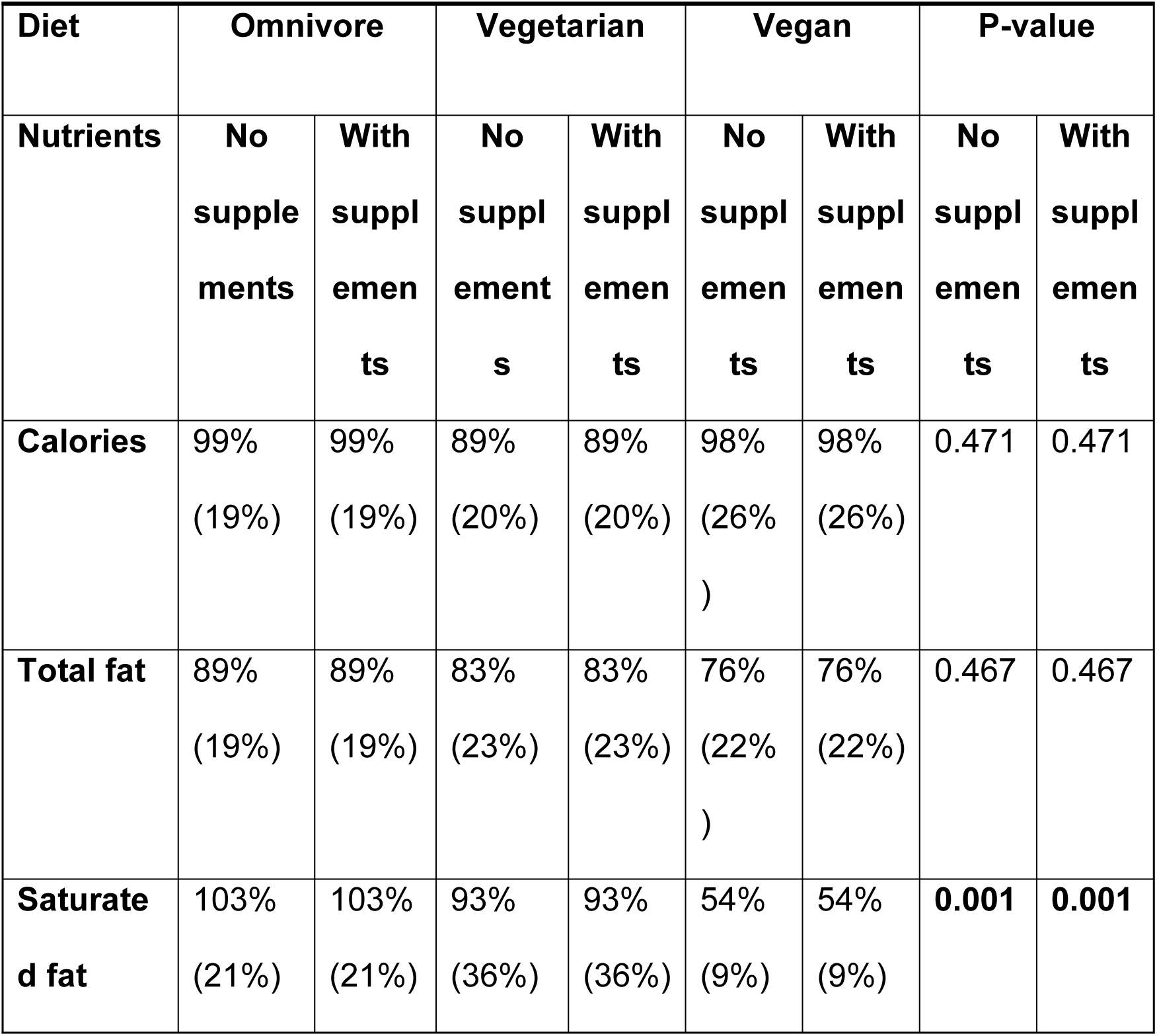
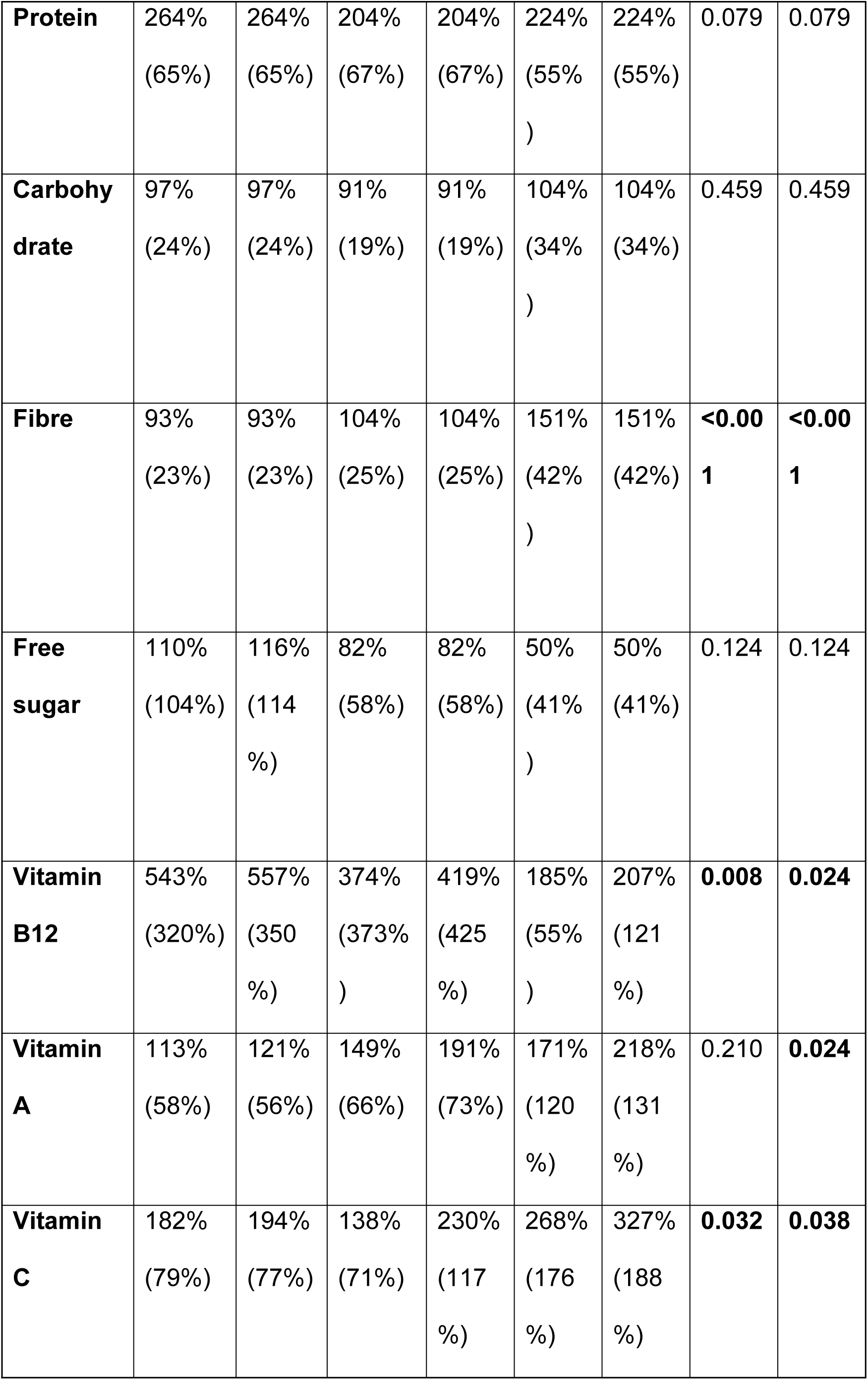
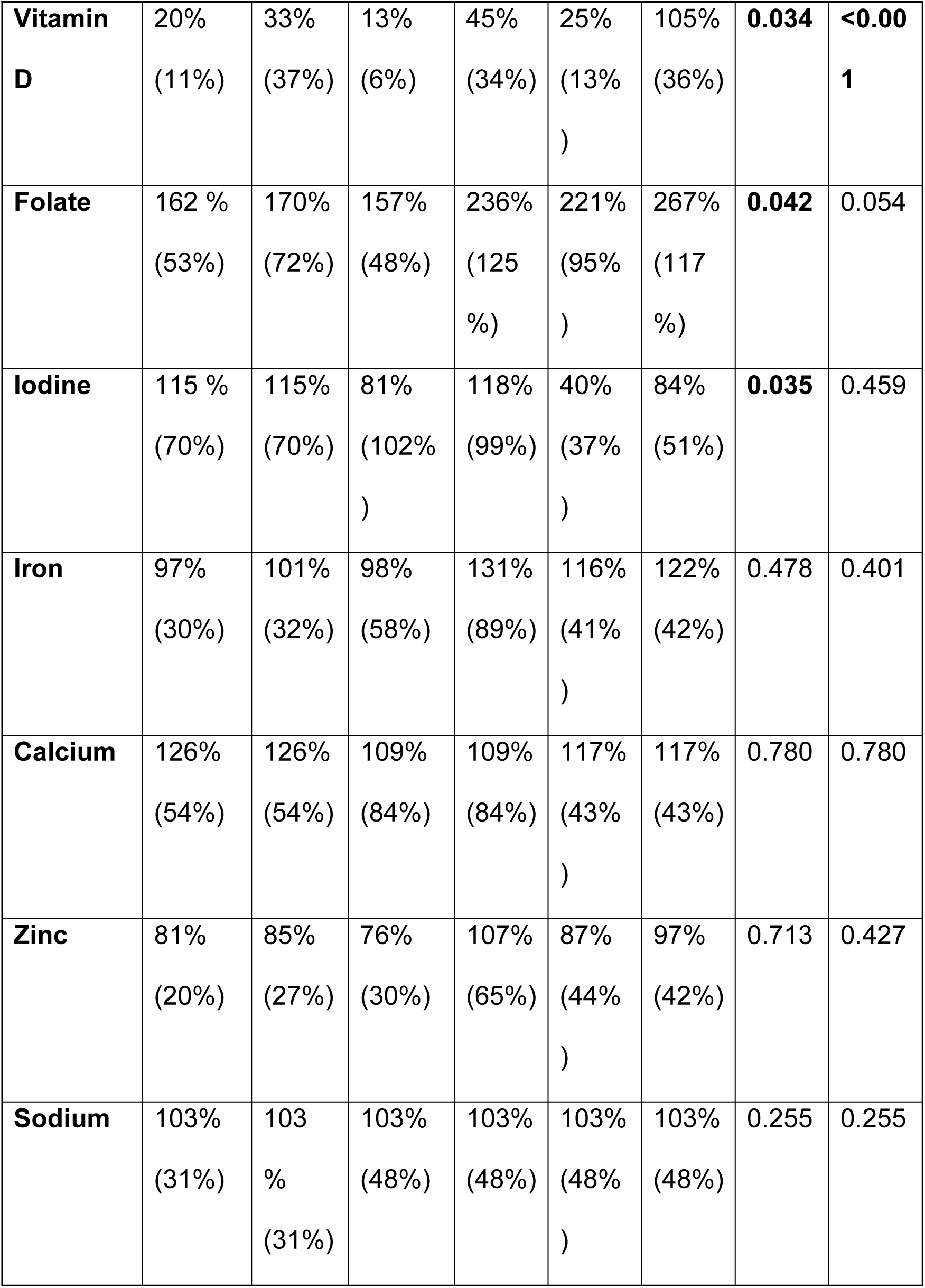
Percentage of nutrient recommendations met per dietary pattern, with and without supplements included, adjusted for age.

| **Diet** | **Omnivore** |  | **Vegetarian** |  | **Vegan** |  | **P-value** |  |
| --- | --- | --- | --- | --- | --- | --- | --- | --- |
| **Nutrients** | **No**<br>**supple**<br>**ments** | **With**<br>**suppl**<br>**emen**<br>**ts** | **No**<br>**suppl**<br>**ement**<br>**s** | **With**<br>**suppl**<br>**emen**<br>**ts** | **No**<br>**suppl**<br>**emen**<br>**ts** | **With**<br>**suppl**<br>**emen**<br>**ts** | **No**<br>**suppl**<br>**emen**<br>**ts** | **With**<br>**suppl**<br>**emen**<br>**ts** |
| **Calories** | 99%<br>(19%) | 99%<br>(19%) | 89%<br>(20%) | 89%<br>(20%) | 98%<br>(26%<br>) | 98%<br>(26%) | 0.471 | 0.471 |
| **Total fat** | 89%<br>(19%) | 89%<br>(19%) | 83%<br>(23%) | 83%<br>(23%) | 76%<br>(22%<br>) | 76%<br>(22%) | 0.467 | 0.467 |
| **Saturate**<br>**d fat** | 103%<br>(21%) | 103%<br>(21%) | 93%<br>(36%) | 93%<br>(36%) | 54%<br>(9%) | 54%<br>(9%) | **0.001** | **0.001** |
| **Protein** | 264%<br>(65%) | 264%<br>(65%) | 204%<br>(67%) | 204%<br>(67%) | 224%<br>(55%)<br>) | 224%<br>(55%) | 0.079 | 0.079 |
| **Carbohydrate** | 97%<br>(24%) | 97%<br>(24%) | 91%<br>(19%) | 91%<br>(19%) | 104%<br>(34%)<br>) | 104%<br>(34%) | 0.459 | 0.459 |
| **Fibre** | 93%<br>(23%) | 93%<br>(23%) | 104%<br>(25%) | 104%<br>(25%) | 151%<br>(42%)<br>) | 151%<br>(42%) | **<0.001** | **<0.001** |
| **Free sugar** | 110%<br>(104%) | 116%<br>(114%) | 82%<br>(58%) | 82%<br>(58%) | 50%<br>(41%)<br>) | 50%<br>(41%) | 0.124 | 0.124 |
| **Vitamin B12** | 543%<br>(320%) | 557%<br>(350%) | 374%<br>(373%) | 419%<br>(425%) | 185%<br>(55%)<br>) | 207%<br>(121%) | **0.008** | **0.024** |
| **Vitamin A** | 113%<br>(58%) | 121%<br>(56%) | 149%<br>(66%) | 191%<br>(73%) | 171%<br>(120%) | 218%<br>(131%) | 0.210 | **0.024** |
| **Vitamin C** | 182%<br>(79%) | 194%<br>(77%) | 138%<br>(71%) | 230%<br>(117%) | 268%<br>(176%) | 327%<br>(188%) | **0.032** | **0.038** |
| **Vitamin D** | 20%<br>(11%) | 33%<br>(37%) | 13%<br>(6%) | 45%<br>(34%) | 25%<br>(13%)<br>) | 105%<br>(36%) | **0.034** | **<0.001** |
| **Folate** | 162 %<br>(53%) | 170%<br>(72%) | 157%<br>(48%) | 236%<br>(125 %) | 221%<br>(95%<br>) | 267%<br>(117 %) | **0.042** | 0.054 |
| **Iodine** | 115 %<br>(70%) | 115%<br>(70%) | 81%<br>(102%<br>) | 118%<br>(99%) | 40%<br>(37%<br>) | 84%<br>(51%) | **0.035** | 0.459 |
| **Iron** | 97%<br>(30%) | 101%<br>(32%) | 98%<br>(58%) | 131%<br>(89%) | 116%<br>(41%<br>) | 122%<br>(42%) | 0.478 | 0.401 |
| **Calcium** | 126%<br>(54%) | 126%<br>(54%) | 109%<br>(84%) | 109%<br>(84%) | 117%<br>(43%<br>) | 117%<br>(43%) | 0.780 | 0.780 |
| **Zinc** | 81%<br>(20%) | 85%<br>(27%) | 76%<br>(30%) | 107%<br>(65%) | 87%<br>(44%<br>) | 97%<br>(42%) | 0.713 | 0.427 |
| **Sodium** | 103%<br>(31%) | 103 %<br>(31%) | 103%<br>(48%) | 103%<br>(48%) | 103%<br>(48%<br>) | 103%<br>(48%) | 0.255 | 0.255 |

Vegetarians do not meet the recommended intakes for calories, total fat, saturated fat, carbohydrate, free sugar, and vitamin D. However, they exceed the recommendations for protein, fibre, vitamins B12, A, and C, sodium, iodine, iron, calcium, zinc and folate when supplements are included. Excluding supplements, there is a decrease in the intake of vitamins B12, A, C, and folate, though these remain above the recommended levels. Iodine, zinc and iron also decrease without supplements, falling below the recommended intake, whereas they meet the recommended levels when supplements are taken into account. Vitamin D decreases further from the recommended intake in the absence of supplements.

Vegans do not meet the recommended intake for calories, total fat and saturated fat, free sugar, iodine and zinc, and exceed the recommended amount for protein, carbohydrate, fibre, vitamin B12, A and C, Sodium, iron, calcium, folate and vitamin D when supplements are included. Excluding supplements, there is a decrease in vitamin B12, A and C, iron, and folate, however, they remain above the recommended levels. Vitamin D also decreases without supplements, falling below the recommended intake, whereas they meet the recommended level when supplements are included. Iodine and zinc decrease further from the recommended intakes in the absence of supplements.

When comparing significant differences between the dietary patterns, ANOVA results indicated an overall significant difference for saturated fat, fibre, vitamins A, C, B12 and D when supplements are included (P values= 0.001, <0.001, 0.024, 0.038, 0.024, <0.001 respectively). Specifically, there was a significant difference between vegans and omnivores for vitamins A, C, B12 and folate, with vegans consuming more vitamins A, C and folate and less B12. For saturated fat, fibre and vitamin D, significant differences were observed between vegans and omnivores, with vegans consuming more fibre and vitamin D and less saturated fat, and vegans and vegetarians with vegan consuming more fibre, vitamin D and less saturated fat, but no significant difference was found between vegetarians and omnivores.

When supplements were excluded, there was an overall significant difference in saturated fat, fibre, vitamins B12, C and D, iodine, and folate (P values= 0.001, <0.001, 0.008, 0.032, 0.034, 0.035, 0.042 respectively). However, upon examining the differences between groups, vitamins C and D showed significant differences only between vegans and vegetarians, with vegans consuming more of both. For iodine, vitamin B12 and folate, a significant difference was found only between vegans and omnivores, with vegans consuming more folate and less iodine and B12.

Table 3 shows the percentage of energy intake from carbohydrates, protein, total fat and saturated fat, alongside government recommendations for these. These values remain the same when supplements are both included and excluded, as macronutrients remained unchanged. All diet types exceed the recommended 50% of energy intake from carbohydrates, with omnivores only slightly exceeding this by 1.8%, vegetarians slightly more by 4.76% and vegans the highest at 6.35% over the recommended intake. However, there are no significant differences between the groups according to the ANVOA analysis. The recommended energy intake for protein is 15%, the only group to meet this is omnivores at 15.44%. Vegetarians and vegans are both slightly under this with vegans reaching 13.2% and vegetarians slightly behind at 12.91% and this is a significant difference between groups. No diets met the recommended energy intake of 35% for fat, with omnivores reaching 32.78%, vegetarians 32.34 and vegans 30.46%, with no significant difference found. However, omnivores and vegetarians exceeded the recommended level of 11% of daily energy intake from saturated fat, 12.37% and 11.18% retrospectively., whereas vegans did not meet this at 7.68%. There was a significant difference seen here between omnivores and vegans and vegetarians and vegans, but not between omnivores and vegetarians.

**Table 3:** Mean nutrient percentage of energy intake by macronutrients per day by dietary pattern.

| **Diet** | **Omnivore** | **Vegetarian** | **Vegan (StDev)** | **P-Value** |
| --- | --- | --- | --- | --- |
| **Nutrients** | **(StDev)** | **(StDev)** |  |  |
| **% of energy from carbohydrate (50% rec)** | 51.8 (4.97) | 54.8 (4.07) | 56.4 (7.37) | 0.124 |
| **% of energy from protein (15%)** | 15.4 (2.16) | 12.9 (2.03) | 13.2 (2.11) | **0.006** |
| **% of energy from fat (35%)** | 32.8 (4.20) | 32.3 (3.64) | 30.5 (7.45) | 0.504 |
| **% of energy from saturated fat (11%)** | 12.4 (2.96) | 11.2 (3.68) | 7.7 (2.25) | **<0.001** |

Table 4 shows the primary food sources of key nutrients among the different dietary groups. Omnivores rely a lot on animal-based products – meat, dairy and eggs. Breakfast cereals in the UK are fortified with various vitamins and are seen to be a main food source of these nutrients for all the dietary patterns, particularly omnivore and vegan. Omnivores do not rely on supplements for any of the nutrients highlighted, whereas these feature within the vegetarian and vegan diets. Fortified dairy alternatives are a main food source for vegans for all the key nutrients, where dairy is for omnivores and vegetarians.

**Table 4:** Primary food sources of key nutrients across omnivore, vegetarian and vegan diets.

|  | **Omnivore** | **Vegetarian** | **Vegan** |
| --- | --- | --- | --- |
| **B12** | Dairy | Dairy | Dairy alternatives |
|  | Beef | Eggs | Nutritional Yeast |
|  | Pork | Nutritional Yeast |  |
| **Iron** | Breakfast cereal | Breakfast cereal | Tofu |
|  | Peas | Bread | Breakfast cereal |
|  |  | Baked beans | Dairy alternatives |
| **Iodine** | Dairy | Supplement | Dairy alternative |
|  | Eggs | Dairy | Supplement |
|  |  | Eggs |  |
| **Vitamin D** | Breakfast cereal | Supplement | Supplement |
|  | Dairy | Eggs | Dairy alternatives |
|  | Pork | Dairy alternatives | Breakfast cereal |
| **Calcium** | Dairy | Dairy | Dairy alternatives |
|  | Breakfast cereal | Bread | Bread |
|  |  | Dairy alternatives |  |
| **Protein** | Dairy | Dairy | Bread |
|  | Chicken | Meat alternatives | Dairy alternatives |
|  | Beef | Eggs | Tofu |

### 3.4 Environmental analysis

Table 5 shows the average daily GHGEs for each dietary pattern, with the significance value from the ANOVA test. Omnivore diets have the largest mean GHGEs (2.53 kgCO₂e), followed by vegetarians (1.70 kgCO₂e) and then vegans (1.36 kgCO₂e). Overall, these differences are statistically significant (P value=<0.001), but post-hoc comparisons revealed that only omnivore diets have a significantly higher carbon footprint than vegan and vegetarian diets, there is no significant difference between vegan and vegetarian diets. This represents a 33% lower footprint for vegetarian diets and a 46% lower footprint for vegan diets relative to omnivores. When comparing vegetarian and vegan diets directly, vegan diets produced 20% fewer emissions.

**Table 5:**
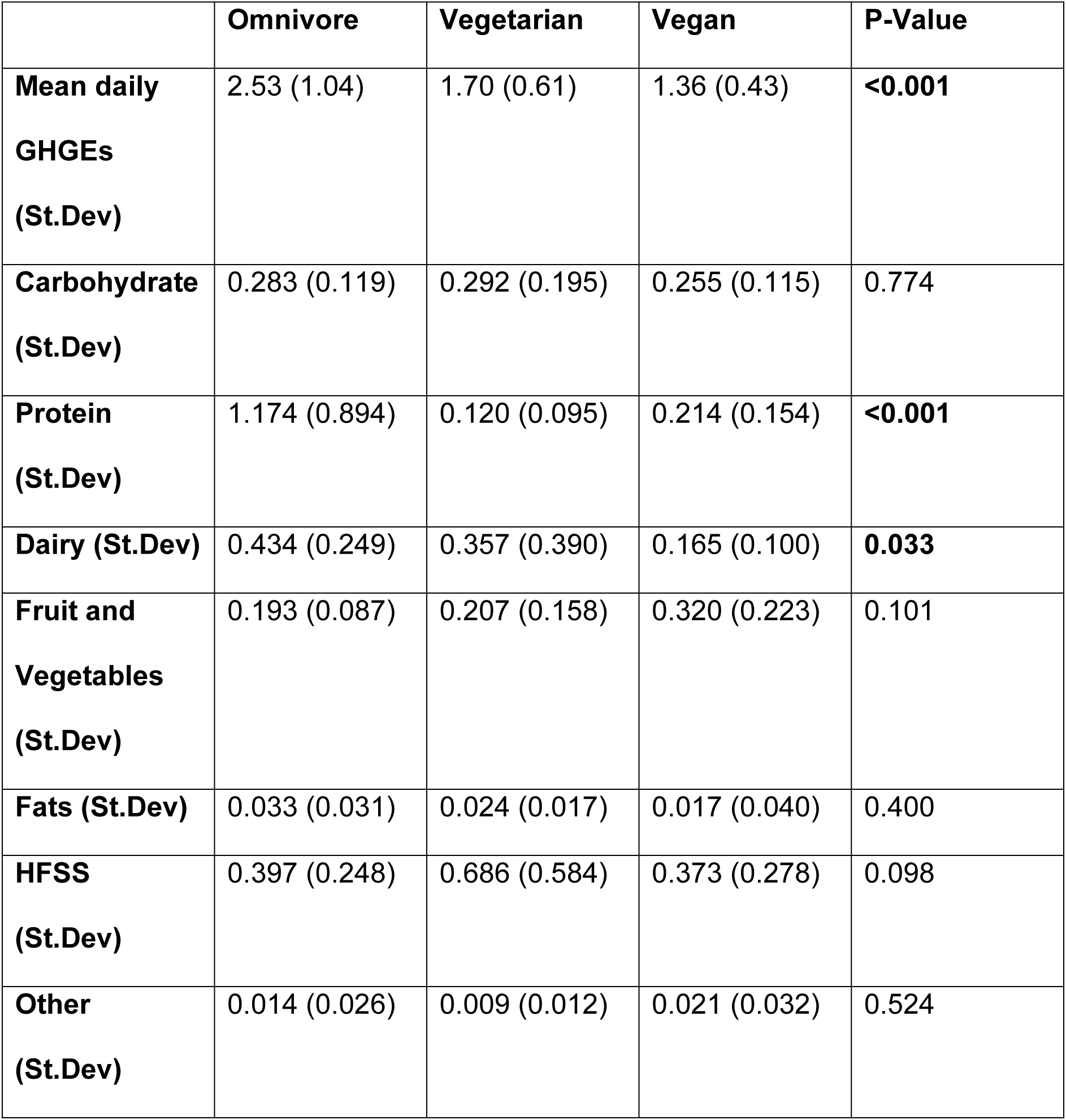
Comparison of mean daily GHGEs (kgCO₂e) across dietary groups and for each Eatwell Guide section, with significance values.

|  | Omnivore | Vegetarian | Vegan | P-Value |
| --- | --- | --- | --- | --- |
| **Mean daily**<br>**GHGEs**<br>**(St.Dev)** | 2.53 (1.04) | 1.70 (0.61) | 1.36 (0.43) | **<0.001** |
| **Carbohydrate**<br>**(St.Dev)** | 0.283 (0.119) | 0.292 (0.195) | 0.255 (0.115) | 0.774 |
| **Protein**<br>**(St.Dev)** | 1.174 (0.894) | 0.120 (0.095) | 0.214 (0.154) | **<0.001** |
| **Dairy (St.Dev)** | 0.434 (0.249) | 0.357 (0.390) | 0.165 (0.100) | **0.033** |
| **Fruit and**<br>**Vegetables**<br>**(St.Dev)** | 0.193 (0.087) | 0.207 (0.158) | 0.320 (0.223) | 0.101 |
| **Fats (St.Dev)** | 0.033 (0.031) | 0.024 (0.017) | 0.017 (0.040) | 0.400 |
| **HFSS**<br>**(St.Dev)** | 0.397 (0.248) | 0.686 (0.584) | 0.373 (0.278) | 0.098 |
| **Other**<br>**(St.Dev)** | 0.014 (0.026) | 0.009 (0.012) | 0.021 (0.032) | 0.524 |

Analysis of mean daily GHGEs by Eatwell Guide section identified significant differences between dietary groups for protein and dairy. For protein, omnivores recorded the highest mean emissions (1.174 kgCO₂e), followed by vegans (0.214 kgCO₂e) and vegetarians (0.120 kgCO₂e). ANOVA confirmed a significant group effect (p < 0.001), with post-hoc comparisons indicating that omnivores differed significantly from both vegetarians and vegans, whereas no significant difference was found between the two PB groups.

For dairy, omnivores recorded the highest mean emissions (0.434 kgCO₂e), followed by vegetarians (0.357 kgCO₂e) and vegans (0.165 kgCO₂e). ANOVA showed a significant effect of dietary group (p = 0.033), with post-hoc tests revealing that vegans had significantly lower dairy-related emissions than omnivores, but no significant difference was found between vegetarians and vegans.

No significant group differences were observed for carbohydrate, fruit and vegetables, fats, HFSS foods, or the ‘other’ category (all p > 0.05). Although vegans had the highest mean GHGEs for fruit and vegetables, ‘other’ omnivores for fats, and vegetarians for HFSS foods and carbohydrates, these differences did not reach statistical significance at the 5% level.

## 4. Discussion

This cross-sectional study assessed the nutritional adequacy and environmental impact of omnivorous, vegetarian, and vegan diets among UK children aged 2–12. The findings reveal that no dietary pattern fully met all nutrient reference values; however, vegetarian and vegan diets were associated with significantly lower GHGEs compared with omnivorous diets. To provide a thorough evaluation, nutrient intakes were analysed both with and without the inclusion of dietary supplements, based on parent-reported data.

Our study finds that children following omnivorous diets exceeded recommended intakes for saturated fat and free sugars while failing to meet the recommended intake for fibre, which mirrors the recent NDNS findings (6), whereas vegetarian and vegan children had intakes of saturated fat, free sugars and fibre in the healthy range. For nutrients commonly associated with animal-derived foods, such as protein and vitamin B12, both vegan and vegetarian diets were found to be adequate even in the absence of supplementation. Vegan children also met iron requirements from diet alone, whereas omnivore and vegetarian children did not meet iron targets without supplementation. Vitamin D intake was insufficient across all groups when supplements were excluded, with only vegan children achieving recommended levels through supplementation. Zinc requirements were met only by vegetarian children with the aid of supplements and were not met by vegan or omnivore children with or without supplementation; while iodine intake remained inadequate in vegan children even with supplementation and was only met by vegetarians when supplements were included, while being met by omnivores with or without supplements.

Energy and total fat intakes were below recommended levels across all groups. Although excessive energy and total fat consumption is associated with increased obesity risk, insufficient intake during childhood is equally concerning, as it may impair growth and development (19). These low intakes are likely due to under reporting, which is a common problem in diet surveys, the most recent NDNS found energy intake was also low in all groups, with under reporting being the likely cause (6). When adjusted for recommended intake, omnivores, vegans, and vegetarians consumed approximately 99%, 98%, and 89% of energy needs, respectively, with no significant group differences. Although total fat intake was low in the omnivore group, they exceeded the recommended daily amount for saturated fat, whereas vegetarian and vegan groups did not. Vegans had a significantly lower intake than vegetarians and omnivores. This suggests that vegans’ fat sources are healthier, which echoes the findings of studies in adulthood (20), this is a positive finding for vegans, as diets high in saturated fat, can lead to increased risks of CVD and stroke (21).

Despite some common concerns about PB protein intake, all dietary groups in this study exceeded protein requirements- by over 200%, showing PB diets can meet protein needs. Nonetheless, when assessing protein as a percentage of total energy intake, only omnivores met the recommended 15%, while vegetarians and vegans fell slightly below this level. This difference was statistically significant. While the total protein quantity was sufficient, it is unknown if all essential amino acids were consumed, as this was not assessed. Previous studies in adults have shown that PB diets may fall short of some essential amino acids (22,23), so further research in children is needed, as it is important they get the essential amino acids for growth and development.

Fibre is a key strength of PB diets, being notably higher in vegans and vegetarians compared with omnivores, and exceeding recommendations. This is similar to previous findings, with PB children consistently consuming more fibre than omnivore children (8,10,24). The recent NDNS results have highlighted fibre as a key concern, with 86% of 4-10 yr olds not meeting the recommendations (6). Fibre plays a crucial role in future health, including reducing the risk of colorectal cancer and improving the gut microbiome (25).

High consumption of free sugars is associated with increased risks of obesity, dental problems, and T2D (26). The incidence of T2D has increased in children, where it was previously associated as an adult health condition, potentially due to increased levels of sugar in the diet (27). In this study, omnivores were the only group to exceed free sugar recommendations at 110%, with vegetarians consuming 82% of the recommended intake and vegans consuming only 50% of the recommended intake. This is similar to what was found in the VeChi study for children and adolescents (8). With the NDNS also finding that children in the UK are consuming far too much free sugar (6).

Vitamin B12 is often cited as a nutrient that could be lacking in PB diets (28,29), and is crucial for children’s brain development and growth (30), so this could be a cause for concern in PB diets. However, in this study, all dietary groups, including vegan met the required intake for vitamin B12, with omnivores consuming the highest quantities and vegans the lowest, with a significant difference seen between vegans and omnivores. Although B12 level did decrease when supplements were excluded, all diets still met the required levels. In PB diets, this was mainly due to fortification of plant milks, with PB children consuming these in large quantities, and nutritional yeast. This highlights the importance of fortifying PB products with micronutrients found in cow’s milk. However, a recent article found that most plant-milks do not contain enough nutrients for a like-for-like swap (31), so it is important there is more fortification across all brands. The VeChi study had different findings, and found vegan children did not have sufficient intake of B12 through diet, but their blood levels remained sufficient (8). This study did not include supplement intake, therefore, although dietary levels of B12 were not sufficient, blood levels being sufficient indicates potential use of supplements. This supports our study findings, suggesting that supplementation and fortification can be effective when used appropriately.

Iodine is another nutrient that has been highlighted as a potential nutrient risk for PB diets (32), which is of particular importance in children for growth and cognitive function (6). In this study, omnivorous and vegetarian children only met their iodine requirements when supplements were included; however, vegan children did not, achieving only 84% of the recommended intake. Although this difference was not statistically significant when supplements were included, it was when supplements were excluded. In the absence of supplementation, iodine intake remained unchanged for omnivores - who generally did not take iodine-containing supplements - but dropped from 118% to 81% in vegetarians and from 84% to just 40% in vegans. The significant difference between vegans and omnivores in the non-supplemented analysis highlights the potential risk of deficiency among vegan children. This is supported in the literature, with the VeChi study highlighting vegan and vegetarian children at risk of deficiency, although they also saw this in omnivore children, unlike this study (8). Although the majority of vegan parents supplied their children with vitamins, these did not contain sufficient quantities of iodine.

Iron intake was highest among vegan children, mainly due to consumption of fortified cereals and tofu, with omnivores having the lowest intake. Without supplements, omnivores and vegetarians fell slightly below recommended levels, 97% for omnivores and 98% for vegetarians. These findings align with some international studies (8), but differ from others (10), likely due to national variance in mandatory food fortification. Calcium intake was also sufficient across all dietary groups in this study, contrasting to what international studies found (8,10), again, this difference is likely due to different mandatory fortification of products with calcium.

Zinc is crucial for children’s growth and development, with low levels having shown a risk of stunting (33). Without supplementation no dietary groups met the recommended zinc intake, with vegetarians having the lowest levels (76%) and vegans having the highest (87%). When supplements were included, vegetarians recorded the highest average zinc intake and were the only group to exceed the recommended value, indicating that zinc supplementation was more common among this group. Vegans fell just short of the recommended intakes with supplements (97%). This is different to what the NDNS reported, as they found the majority of children were meeting the recommended intake for zinc (6).

Vitamin D emerged as a critical nutrient of concern. Consistent with national data from the NDNS (6), this study found that vitamin D intake was below recommended levels in all groups, regardless of dietary pattern. Only the vegan group met the recommended intake when supplements were included, reflecting a higher overall rate of supplementation in this group. When supplements were excluded, none of the groups met the reference intake. This is particularly concerning, as vitamin D is essential for calcium absorption and bone development, and deficiency in childhood can lead to long-term skeletal issues (34). Vitamin D supplementation is recommended in the UK in Autumn and winter. As this study was conducted between March and November, this may be why the majority of participants did not supplement this, and blood levels of vitamin D might indicate sufficient levels.

From an environmental perspective, this study is the first to assess and compare the carbon footprints of omnivorous, vegetarian, and vegan diets specifically in UK children. The findings align with the well-established literature on adult populations: omnivorous diets had the highest GHGEs, followed by vegetarian and then vegan diets (35,36). The difference between omnivorous and PB groups was statistically significant, although no significant difference was observed between vegetarian and vegan diets. The observed differences in carbon footprints between dietary groups align with the current literature, reflecting the environmental burden of specific food groups. Protein-related emissions were significantly higher among omnivores, consistent with evidence that meat is among the most emissions-intensive foods due to livestock production and feed requirements (37,38). By contrast, both vegetarians and vegans showed substantially lower protein-related carbon footprints, highlighting the environmental benefit of reducing reliance on animal protein sources. Dairy-related emissions were also highest among omnivores, followed by vegetarians, with vegans recording the lowest values. The difference was significant between omnivores and vegans but not between omnivores and vegetarians, suggesting that vegetarian diets may involve greater reliance on dairy to replace meat, as also observed in adult populations (39). The exclusion of dairy by vegans resulted in the lowest emissions, further supporting evidence that PB alternatives offer carbon benefits (40). These results emphasise the environmental burden of dairy and point towards the potential role of fortified PB alternatives in reducing emissions while maintaining nutritional adequacy in children. No significant differences were observed across other Eatwell Guide sections, including carbohydrate, fruit and vegetables, fats, HFSS foods, or the ‘other’ category, suggesting that diet group differences are most pronounced in relation to animal protein and dairy consumption. These findings reinforce the evidence that dietary transitions away from animal-based protein and dairy represent the greatest potential for reducing the carbon footprint of children’s diets.

This study is the first to explore both the nutritional adequacy and carbon footprints of PB diets in UK children, providing novel insights into a previously unexamined population. However, several limitations were highlighted.

Firstly, the small sample size and method of recruiting participants limits the statistical power and generalisability of the results. While this study found protein intake was adequate in PB diets, the specific amino acid intakes were not explored, this would be beneficial to understand to ensure PB children consume all essential amino acids. Energy intake and total fat intake were lower than expected, which is likely due to under reporting by participants, a common limitation found in dietary studies. The use of convenience sampling meant that the study population were less diverse than the general population and not representative, limiting the generalisability of the results. Another limitation is the analysis of the environmental impact of the diets, this was only assessed using carbon footprints and did not take other environmental factors into account, such as water-usage or packaging.

Dietary supplements were excluded from the carbon footprint calculations due to their minimal intake (approximately 1g/day among users), however it is unlikely that their inclusion would have meaningfully altered the overall results. Previous research has shown that the largest contributors to the environmental impact of supplements stem from packaging and transportation, particularly when plastic containers or blister packs are used (41). However, as packaging details were not captured in the food diaries, this study did not attempt to estimate these effects. Supplement usage varied considerably across dietary groups, with only 13% of omnivores taking supplements compared to 82% of vegetarians and 92% of vegans. The likely reason for the PB participants to take more supplements, is a potential concern of deficiencies through exclusion of animal products from the diet. As mentioned, animal products are often associated with higher environmental impacts compared to PB products, therefore, nutritional supplements, consumed in such low quantities will likely have a far lower environmental impact than consuming animal products.

A strength of this study was that it was the first of its kind within a UK setting, where both vegetarian and vegan intakes were observed to assess nutrient intakes and carbon footprints. The use of three-day weighed food diaries offers greater accuracy compared with more commonly used dietary assessment tools such as food frequency questionnaires or 24-hour recalls. Another strength is the dual analysis of nutrient intake both including and excluding supplements, which provides a more comprehensive picture of dietary sufficiency. Additionally, the study’s environmental analysis, conducted at the ingredient level, offers new insights into the carbon footprints of children’s diets - an area that has been largely overlooked in the literature.

This study does highlight areas for future research. Larger studies are needed, with more representative cohorts to understand if these results are replicated in larger populations and longitudinal research is needed to assess long-term health impacts of PB diets in childhood. Future studies analysing the amino acid profiles of the dietary groups is needed, particularly for PB diets, to understand if these children are consuming the right protein profiles. Studies should also be conducted where blood concentrations of nutrients are taken, as the dietary intake in this study was assessed over 3 days which may not be typical of routine dietary behaviour, whereas blood concentrations would give a good picture of general dietary intakes. In addition, blood concentrations give more information about bioavailability, for example iron may be more readily absorbed into the bloodstream if it is sourced from animals, so that even though vegan and vegetarian children had as much or more dietary iron as omnivores, it is possible that this does not translate into blood concentration. Finally, although more PB diets could be recommended, the availability and affordability of these around the UK needs to be known, to ensure that any policy changes are equitable for all and do not further increase health inequalities.

## 5. Conclusion

This study is the first to evaluate both the nutritional adequacy and carbon footprints of omnivorous, vegetarian, and vegan diets in UK children aged 2–12. The findings suggest that while no dietary pattern met all nutrient recommendations, well-planned PB diets, particularly when supported by fortified foods and appropriate supplementation, can provide adequate nutrition for children. Vegan and vegetarian diets were associated with higher intakes of fibre, folate, and vitamins A and C, and lower intakes of saturated fat and free sugars compared with omnivorous diets. However, nutrients such as iodine, vitamin D, and zinc emerged as areas of concern in PB diets, particularly if supplements are not used.

From an environmental perspective, omnivorous diets had significantly higher carbon footprints than vegetarian and vegan diets, confirming that dietary shifts towards PB eating could play a meaningful role in reducing GHGEs, even in children. These findings reinforce the potential for dietary guidelines that consider both health and sustainability.

Given the rising interest in PB diets in the UK, particularly in young adults, who may be raising PB children in the future, these results support the development of guidelines to support this group, as these would promote child and planetary health. In this study, all vegan children had packed lunches (even those within the age-groups that would receive free school meals), indicating potentially poor PB choices in school. Changes could be made in school food standards, shifting them to being more PB than current standards to accommodate these children in the future. More mandatory fortification of products, particularly those replacing animal-based products where nutrients may be lacking, could also support nutritional adequacy in PB children.

## Data Availability

Data cannot be shared publicly because no consent was given by the participants to openly share their data, where data was to only be accessed by study researchers. Data are available from the main researchers (Alice Coffey: or Robert Lillywhite), as in line with the University of Warwick's Biomedical and Scientific Research Ethics Committee (contact via) for researchers who meet the criteria for access to confidential data.

